# LIFESTYLE CHANGES IN PATIENTS WITH NON-ALCOHOLIC FATTY LIVER DISEASE: A SYSTEMATIC REVIEW PROTOCOL

**DOI:** 10.1101/2020.06.22.20137430

**Authors:** Tiziana Fernández-Mincone, Macarena Viñuela Morales, Marco Arrese Jiménez, Juan Pablo Arab Verdugo, Daniel Cabrera, Francisco Barrera Martínez

## Abstract

Non-alcoholic fatty liver disease (NAFLD) is a worldwide raising liver condition and it is expected to become the first cause of progression to cirrhosis and hepatocellular carcinoma in the next 5 years (Araújo 2018). At present, there are no pharmacological treatments approved. Weight loss is the first-line treatment (European Association for the Study of the Liver 2016) (Chalasani 2012) showing that 7 to 10% weight loss improves steatosis, inflammation, hepatocyte ballooning and, in NASH patients, has an impact in fibrosis too (Romero-Gómez 2017). However, the reduction and maintenance of weight loss is often complex. To achieve this goal, it is recommended doing lifestyle changes including exercise and diet on daily living (Hannah 2016). However, it has been difficult to establish any conclusions regarding the best type of diet or exercise protocol to maximize improvements and to maintain its effects through time. Because of this, the objective of this review is assessing the efficacy and safety of diet, exercise, or a combination of these in patients with NAFLD.

## Introduction

Non-alcoholic fatty liver disease (NAFLD) is a condition that includes a spectrum of liver diseases ranging from simple steatosis, entity where lipid droplets accumulates within the cytoplasm in more than 5% of hepatocytes, to non-alcoholic steatohepatitis (NASH), characterized by steatosis, inflammation and hepatocyte cell ballooning associated with fibrosis (Arab 2018) (Malaguarnera 2009), not caused by other factors. It is strongly associated with obesity, insulin resistance (IR), type II Diabetes Mellitus (T2DM) and dyslipidemia (EASL-EASD-EASO 2016).

Its prevalence is 24-25% in general population and it is expected to become the first cause of progression to cirrhosis and hepatocellular carcinoma in the next 5 years (Araújo 2018).

Weight loss is the first-line treatment of non-alcoholic liver disease (European Association for the Study of the Liver 2016) (Chalasani 2012). A 7 to 10% weight loss improves steatosis, inflammation, hepatocyte ballooning and, in NASH patients, has an impact in fibrosis too (Romero-Gómez 2017). However, the reduction and maintenance of weight loss is often complex

The strategies to achieve this goal are lifestyle changes which includes exercise and diet therapy (Hannah 2016). Exercise has a large effect in intrahepatic triglycerides, enzymes levels, insulin sensitivity, glucose homeostasis (Katsagoni 2017) (Kenneally 2017), free fat acids transportation and oxidation, radical of oxygen production and inflammation (van der Windt 2018). Current guidelines for management of NAFLD recommends 150 minutes per week of moderate exercise (Leoni 2018), similar to what American College of Sports Medicine indicates. About diet, it has been suggested that decreasing caloric intake between 750-1000 kcal/day or 30% of the intake, improves IR and hepatic steatosis (Chalasani 2018) and its composition can reduce free fatty acids, fasting glucose, IR and alanine-transferase (Dongiovanni 2016). In combination, diet plus exercise have demonstrated improvements in liver histology, liver enzymes (Katzagoni 2017) and weight reductions of 4.2% to 10.6% (Thoma 2012).

There are several publications about lifestyle changes and NAFLD reduction (Wong 2013) (Younossi 2017) (Zou 2018). However, it has been difficult to establish any conclusions regarding the best type of diet or exercise protocol to maximize improvements and to maintain its effects through time.

### Objectives

To assess the efficacy and safety of diet, exercise, or a combination of these in patients with NAFLD.

## Methods

### Criteria for considering studies for this review

#### Types of studies

we will only include randomized clinical trials. Studies evaluating the effects on animal models or in vitro conditions will be excluded.

#### Types of participants

We will include trials assessing participants older than 18 years old with NAFLD, NASH or non-alcoholic cirrhosis. NAFLD should have been diagnosed based on abnormalities in laboratory test (Kemp 2008), liver biopsy, ecography, ultrasound (US) or magnetic resonance imaging (MRI). NASH and cirrhosis should have been diagnosed by liver biopsy or blood test or scores such as NAFLD fibrosis score or NAFLD activity score as international guidelines mentioned (Leoni 2018).

Studies that consider healthy volunteers, patients with other liver conditions such as hepatitis, vascular disorders, drugs or toxin induced liver injuries or alcoholic liver disease (Reisner 2015), metabolic syndrome, obesity or T2DM without NAFLD will be excluded.

#### Types of interventions

We will include any trial assessing the effect of lifestyle interventions, considering dietary and/or exercise interventions.

Dietary interventions are defined as any diet with calorie restriction, low fat or low carbohydrates composition. Exercise was defined by Caspersen (Caspersen 1985) as “physical activity that is planned, structured, repetitive, and purposive in the sense that improvement or maintenance of one or more components of physical fitness is an objective”. Physical fitness includes health-related components like cardiorespiratory endurance, muscular endurance, muscular strength, body composition, and flexibility (Caspersen 1985).

The comparison of interest will be conventional/standard/optimal treatment or no treatment, as defined by the authors of the trials.

#### Types of outcome measures

We will not use outcomes as an inclusion criterion during the selection process. Any article meeting all the criteria except for the outcome criterion will be preliminarily included and assessed in full text.

a. **Primary outcomes:**
  - Cardiovascular events
  - All-cause mortality
  - Quality of life
b. **Secondary outcomes**
  - Anthropometric measurements: lean body mass, BMI
  - Physical fitness: maximum/peak oxygen
c. **Other outcomes**
  - Reversion of histological alterations such as steatosis, inflammation, fibrosis, liver cell injury
  - Intrahepatic triglycerides
  - Liver enzymes concentrations
  - HbA1c, HOMA-IR, fasting glucose
  - Lipid profile

### Search methods for identification of studies

#### Electronic searches

We will search the Cochrane HepatolJBiliary Group Controlled Trials Register, the Cochrane Central Register of Controlled Trials (CENTRAL) in the Cochrane Library (latest issue), MEDLINE Ovid, Embase Ovid, LILACS, without any language or publication status restriction. The searches will cover from the inception date of each database until the day before submission.

#### Other sources

In order to identify articles that might have been missed in the electronic searches, we will search for primary studies in the following sources:

- Systematic reviews about NAFLD and lifestyle changes
- Online trial registries such as ClinicalTrials.gov (clinicaltrials.gov) and WHO International Clinical Trial Registry Platform (www.who.int/ictrp) for ongoing or unpublished trials.
- Google Scholar and Microsoft Academic for cross-citation search using each included study as the index reference.
- Reference lists of included studies to identify additional trials for inclusion.

### Data collection and analysis

#### Selection of studies

Two authors (TF and MV) will independently review studies through two stages: 1) title and abstract screening; 2) fulllJtext screening. If they encounter any inclusion or exclusion discrepancies, the two authors will either resolve them by discussion, or consult a third author who will act as arbitrator. We will exclude duplicate studies.

#### Extraction and management of data

Using standardized forms, two reviewers will extract data independently from each included study. We will collect the following information: study design, setting, participant characteristics (age, sex, body mass index, homeostasis model assessment-HOMA-, enzymatic levels, intrahepatic triglycerides levels, NAFLD stage, comorbidities), and study eligibility criteria; details about the intervention and comparison, such as calorie restriction, diet composition and duration in dietary intervention and exercise modality, intensity, frequency and duration in exercise protocol; the outcomes assessed and the time they were measured; losses to follow up, exclusions and reasons; the source of funding of the study and the conflicts of interest disclosed by the investigators; the risk of bias assessment for each individual study.

We will resolve disagreements by discussion, and one arbiter will adjudicate unresolved disagreements

#### Assessment of risk of bias in included studies

Two review authors (TF and MV) will independently assessed the risk of bias using the Cochrane Collaboration’s tool for assessing risk of bias, which includes the following items: random sequence generation, allocation concealment, blinding of participants and personnel, blinding of outcome assessment, incomplete outcome data, selective reporting and other sources of bias. The risk of bias will be classified as high, low or unclear, according to the methods described in Chapter 8 of the Cochrane Handbook for Systematic Reviews of Interventions (Higgins 2011b). Disagreements will be resolved by discussion and consensus.

### Measures of treatment effect

For dichotomous outcomes, we will express the estimate of treatment effect of an intervention as risk ratios (RR) or odds ratios (OR) along with 95% confidence intervals (CI).

For continuous outcomes we will use mean difference and standard deviation (SD) to summarize the data using a 95% CI. Whenever continuous outcomes are measured using different scales, the treatment effect will be expressed as a standardized mean difference (SMD) with 95% CI. For outcomes measure as scores, we will use mean difference and standard deviation. If studies use different scores, the effect will be expressed as standardized mean difference (SMD).

#### Dealing with missing data

We will write to authors of included trials to request additional data as required. If we do not receive a response from the authors of the studies not reporting primary outcomes, we will analyze available data. We will address the potential impact of missing data on the findings of the review in the discussion using worst-case and best-case scenario.

#### Assessment of heterogeneity

We will quantify heterogeneity using I^2^ test, as it does not inherently depend on the number of studies considered. We will present the I^2^ and corresponding 95% CI to gauge the degree of heterogeneity present in the sample.

### Data synthesis

Considering that studies will include either a dietary component in addition to exercise, or different types of exercise or diets, we will restrict inclusion to trials compare lifestyle changes to standard care, optimal, conventional care or no intervention.

For any outcomes where data were insufficient to calculate an effect estimate, a narrative synthesis will be presented, describing the studies in terms of direction and size of the effects, and any available measure of precision. For any outcomes where data is available from more than one trial we will conduct a formal quantitative synthesis (meta-analysis) for studies clinically homogeneous using RevMan 5, using the inverse variance method with the random-effects model.

#### Subgroup analysis

We plan to undertake the following subgroup analyses:

- Intervention category (diet, exercise or diet plus exercise)
- NAFLD stages
- Comorbidities (pure NAFLD versus NAFLD plus comorbidities)
- Type of exercise (resistance, endurance or combination)
- Type of diet
- Exercise intensity (low, moderate or high)

In case we identify significant differences between subgroups (test for interaction <0.05) we report the results of individual subgroups separately.

#### Sensitivity analysis

We will perform sensitivity analysis excluding high risk of bias studies. In cases where the primary analysis effect estimates and the sensitivity analysis effect estimates significantly differ, we will either present the low risk of bias — adjusted sensitivity analysis estimates — or present the primary analysis estimates but downgrading the certainty of the evidence because of risk of bias.

## Data Availability

All the information included in this manuscript is referenced in previous publications

## Authors’ contributions

TF conceived the protocol. TF drafted the manuscript, and all other authors contributed to it. The corresponding author is the guarantor and declares that all authors meet authorship criteria and that no other authors meeting the criteria have been omitted.

